# A review of behavioural rating scales for the assessment of Attention Deficit/Hyperactivity Disorder among adults in research

**DOI:** 10.1101/2023.05.30.23290708

**Authors:** Marius Grandjean, Shachar Hochman, Raja Mukherjee, Roi Cohen Kadosh

**Author notes:** Correspondence: Dr Shachar Hochman.

## Abstract

**Objective:** This review aims to provide researchers with a contemporary and comprehensive understanding of the current state of behavioural rating scales used in evaluating adult ADHD for research purposes. The objective is to offer guidance that enables researchers to make informed decisions when selecting the most suitable scale for their studies. Moreover, our intention was to map and compare these scales, with a specific focus on detecting feigned or invalid symptom presentation—an aspect notably overlooked in prior reviews.

**Method:** We reviewed the most recent literature on behavioural rating scales for adult ADHD assessment. We evaluated the scales and compared them based on their psychometric properties and the range of symptoms that they assessed.

**Results:** The Conners’ Adult ADHD Rating Scales (CAARS), Mind Excessively Wandering Scale (MEWS), and Wender Utah Rating Scale (WURS) have emerged as the most accurate measures for assessing adult ADHD. Moreover, there is an increasing emphasis on the development of assessment tools, either integrated within existing scales or as independent measures, to evaluate feigning or invalid symptom presentation. In that regard, stand-alone measures have demonstrated greater effectiveness compared to embedded measures, with the ADHD Symptom Infrequency Scale (ASIS) being identified as the most accurate scale for the detection of feigning.

**Conclusion:** Based on this review, we provide recommendations for the behavioural rating scales with the most accurate measurement of relevant variables in research-related settings.

## Introduction

Attention Deficit Hyperactivity Disorder (ADHD) is a neurodevelopmental disorder exhibiting clinical heterogeneity, marked by the presence of a sustained tendency towards inattention and distractibility, as well as hyperactivity and impulsivity, causing interference in functioning or development (American Psychiatric Association, 2022). In many cases, ADHD continues to affect individuals into adulthood, and can be associated with an increased risk of developing other mental health disorders, as well as negative outcomes such as educational underachievement, challenges with employment and interpersonal relationships, and potential involvement with criminal activities (Sayal et al., 2018). The phenomenon of adult ADHD can be differentiated into two distinct subtypes, namely persistent and symptomatic adult ADHD. The former subtype is characterised by the enduring presence of ADHD symptoms over an extended period, spanning from childhood through adulthood. Conversely, the latter subtype refers to the presence of clinically significant symptoms of ADHD that cause impairment in daily functioning without a specific childhood-onset (Song et al., 2021). Recent research has reported the global prevalence rates of persistent and symptomatic adult ADHD to be 2.58% and 6.76% respectively, corresponding to 139.84 million and 366.33 million of affected adults worldwide (Song et al., 2021).

When assessing adult ADHD, various methods are used, including interviews, continuous performance tests (CPTs), and behavioural rating scales. CPTs are computer-based neuropsychological tests that require the individual to respond to a series of stimuli, measuring specific cognitive functions that are impaired in ADHD. Meanwhile, behavioural rating scales are subjective measures completed by either the individual or an informant. These scales usually assess the frequency and severity of ADHD-related behaviours. While clinical interviews have often been regarded as the gold standard for ADHD assessment (Murphy & Gordon, 1998; Ramsay, 2015), a recent comprehensive review by Marshall et al. (2021) found that a combination of interviews, CPTs and behavioural rating scales result in the highest rate of diagnostic accuracy. Unfortunately, such an extensive intervention may not always be feasible (Kessler et al., 2005), and it is resource intensive. In research settings, behavioural rating scales and/or CPTs are often used as the primary outcome of studies on new treatment for ADHD (Allenby et al., 2018; Alyagon et al., 2020; Berger et al., 2021; McGough et al., 2019; Nahum et al., 2023; Paz et al., 2018; Weaver et al., 2012). While several studies have examined the effectiveness of CPTs (Baggio et al., 2020; Epstein et al., 2003; Ogundele et al., 2011; Riccio & Reynolds, 2001; Vaughn et al., 2011), only a few have assessed the usefulness of behavioural rating scales (Harrison & Edwards, 2023; Marshall et al., 2021; Taylor et al., 2011). Given their relatively low cost and quick administration, rating scales are often favoured as the main outcome measure in studies focused on new treatments for ADHD (Alyagon et al., 2020; McGough et al., 2019; Paz et al., 2018; Weaver et al., 2012). However, there is a relative lack of a clear understanding of the quality and adequacy of these rating scales. This prevents researchers to make informed decisions when selecting which scale to use and for what purpose.

To enhance the reader’s understanding of the commonly used behavioural rating scales used in the assessment of adult ADHD, we will provide a brief and non-exhaustive overview of the most frequently used measures for assessing adult ADHD. These rating scales are either available publicly at no cost or commercially. Among the publicly available scales, there is the Wender Utah Rating Scale (WURS) which is a retrospective self-reported measure consisting of 61 items that assess experienced childhood symptoms of ADHD in adults. A shorter version of the WURS known as WURS-25 is also available (Ward et al., 1993). Another widely used measure is the Adult ADHD Self-Report Scales (ASRS), which assesses the 18 symptoms of ADHD outlined in the DSM-4 (Adler et al., 2006). In addition to the free measures previously mentioned, there are also commercially available measures. To begin, Barkley Adult ADHD Rating Scale (BAARS-IV) has various subscales for assessing current and childhood symptoms based on DSM-5 criteria. Each scale consists of 27 items and can be administered either through self-report or by an observer (Barkley, 2011). Then, there is the Brown Attention-Deficit Disorder Scales (BADDS), which is a self-reported questionnaire consisting of 40 to 50 items divided into six subscales. It assesses symptoms associated with ADHD and executive function impairments (Brown, 1996), as such impairments has been suggested to be one of the underlying mechanisms of ADHD (Barkley, 1997). Lastly, the Conners’ Adult ADHD Rating Scales (CAARS, long version) has 66 items that assess the presence and severity of ADHD symptoms through statements about daily activities and behavioural tendencies. Similarly, to the BAARS-IV, the CAARS also has both self- and observer-reported versions that comprise identical items.

Interestingly, The CAARS also includes an Inconsistency Index, which is a measure for careless or random responding (Conners et al., 1999). In addition to the original Inconsistency Index, researchers have developed two additional indexes for the CAARS aiming to support the psychometric value of the CAARS; The first is the CAARS Infrequency Index (CII), which consists of items from the CAARS that are rarely endorsed by individuals with or without ADHD. The CII is useful for identifying overreporting of symptoms (J. A. Suhr et al., 2011). The second Index Is the Exaggeration Index (EI), which includes items from the Dissociative Experiences Scale (DES) that are integrated into the CAARS. The EI measures the extent to which an individual reports exaggerated or extreme symptoms of ADHD (Harrison & Armstrong, 2016).

In previous reviews, the CAARS was argumentatively recommended as the most useful measure due to its psychometric properties (Taylor et al., 2011), its coverage of a large constellation of symptoms and its ability to identify invalid symptom presentation (Marshall et al., 2021), which is becoming an increasingly concerning issue. Over the last decade, studies have shown that behavioural rating scales can be easily falsified (Jachimowicz & Geiselman, 2004; Lee Booksh et al., 2010; Quinn, 2003). This issue is compounded by the fact that some individuals may feign ADHD symptoms for a variety of reasons, such as to provide a socially acceptable excuse for their difficulties (J. Suhr & Wei, 2013) or to obtain benefits associated with ADHD medication (Hinshaw & Scheffler, 2014). This is particularly concerning because the non-specific and subjective nature of ADHD symptoms makes it easier for individuals to feign symptoms during formal evaluations, which is especially prevalent among adults, particularly college students. Additionally, the diagnostic criteria for adult ADHD, as emphasised by the DSM-5, rely heavily on subjective symptoms rather than cognitive or functional deficits. Therefore, when using behavioural rating scales to assess ADHD symptoms, it is crucial to consider the possibility of feigned results. The primary objective of this paper is to scrutinize the latest advancements concerning the utilisation of behavioural rating scales in the evaluation of ADHD symptoms. More specifically, this review aims to provide researchers with an updated and comprehensive understanding of the current state of behavioural rating scales utilized in the assessment of adult ADHD, to aid them in making informed decisions when selecting an appropriate scale for their research (for a comprehensive analysis of the diagnostic efficacy of behavioural rating scales for clinical utility, see Harrison & Edwards, 2023).

The traditional approach to compare rating scales used for ADHD assessment is by examining their psychometric properties. However, there is no agreed-upon gold standard regarding which statistics are more important or how to determine when one scale is better than another. In the present review, we chose to focus on evaluating the sensitivity, specificity, PPV, and NPV of the rating scales. Consequently, before delving further into this review, we briefly describe the fundamental statistics that we used. To begin, *sensitivity*, also referred to as the true positive rate, reflects the likelihood that a test or measurement accurately identifies the presence of a specific condition. To elucidate, an ADHD rating scale with a sensitivity of 0.60 would correctly identify ADHD in 60% of cases. Moving on, *specificity*, known as the true negative rate, signifies the probability of a test or measurement accurately recognizing the absence of a particular condition. For instance, an ADHD rating scale boasting a specificity of 0.30 would accurately identify the absence of ADHD in 30% of cases. Transitioning to *positive predictive value* (PPV), which gauges the probability that an individual possesses a condition if the test or measurement identifies them as having it. For example, an ADHD rating scale with a PPV of 0.80 signifies that if the test indicates ADHD presence, there’s an 80% likelihood the individual indeed has ADHD. In contrast, *negative predictive value* (NPV) assesses the probability that an individual lacks a given condition if the test or measurement designates them as not having it. To provide an example, an ADHD rating scale with an NPV of 0.70 would imply a 70% probability of an individual not having ADHD if the test indicates so (Ivnik et al., 2001).

In simpler terms, *sensitivity* measures how well a test can correctly find positive cases. On the other hand, PPV looks at how likely it is that a positive test result truly means the person has the condition. This way, for example, it is possible to have a test that does not catch all positive cases (low *sensitivity*), but when it does yield a positive result, it is usually correct (high PPV). In contrast, *specificity* measures how well the test can correctly identify those without the condition (negative cases). The NPV, meanwhile, gauges how likely it is that a negative test result truly means the person doesn’t have the condition. Some authors argued that although sensitivity and specificity are crucial in determining true positive and true negative, they fail to account for the confidence level a test score can provide in either confirming or excluding a diagnosis or condition. Likewise, PPV and NPV are useful in clinical decision-making because they are influenced by the base rate (BR), which is the prevalence of the condition in the population but do not indicate the frequency with which the test identifies the targeted behaviour. Hence, it is strongly recommended to interpret all four values concurrently and use qualitative descriptors to interpret their clinical usefulness (Lange & Lippa, 2017) (see Table 1).

**Table 1.** Recommended qualitative descriptors for sensitivity, specificity, PPV and NPV values as proposed by Lange & Lippa (2017).

| Value as a percentage | Qualitative descriptor |
| --- | --- |
| <10 | Very low |
| 10-24 | Low |
| 25-39 | Low-moderate |
| 40-59 | Moderate |
| 60-74 | Moderate-high |
| 75-89 | High |
| 90-100 | Very-high |

*Note.* PPV = positive predictive value, NPV = negative predictive value.

Although behavioural rating scales should not be relied upon solely to clinically diagnose adult ADHD (Harrison & Edwards, 2023), they are often regarded as a valuable tool in research settings. This is particularly notable when the objective is to measure changes in symptoms during the implementation of new treatments, prioritizing symptom assessment over establishing a conclusive diagnosis. This situation often prompts researchers to deliberate on the most appropriate scale that aligns with their specific research objectives. This paper sets out to elucidate the current status of these scales, offering in-depth insights into their accuracy, the symptoms they effectively capture, and their distinct characteristics. Through practical recommendations, this paper aims to empower researchers to make informed decisions when selecting the most suitable scale for their research endeavours.

## Materials and Methods

We conducted a comprehensive search in the PubMed database between 2019 and February 2023, using the search terms “ADHD” AND “attention deficit hyperactivity disorder” AND “assessment or testing or evaluation” AND “adult” AND “diagnosis” (see Figure 1). We specifically chose these terms to match those used in the most recent review of adult ADHD assessment (Marshall et al., 2021) which examined the literature published between 1998 and 2019. Therefore, we selected 2019 as our starting point to provide an updated view of the state of behavioural rating scales for adult ADHD assessment. Electronic publications were the only publications considered for inclusion in the review. In the initial search, a total of 1264 abstracts of journal articles were identified, from which 73 were considered potentially relevant to adult ADHD assessment. After a further consideration of the abstracts, the full texts of 51 journal articles were reviewed, in addition to their bibliographies and citations, which led to the identification and review of an additional 5 articles. The final phase of the literature search aimed at identifying articles that met the inclusion criteria culminating in a final sample of 11 articles (see Table 2). A detailed description of the reviewed papers can be found in Appendix 1.

**Figure 1.**
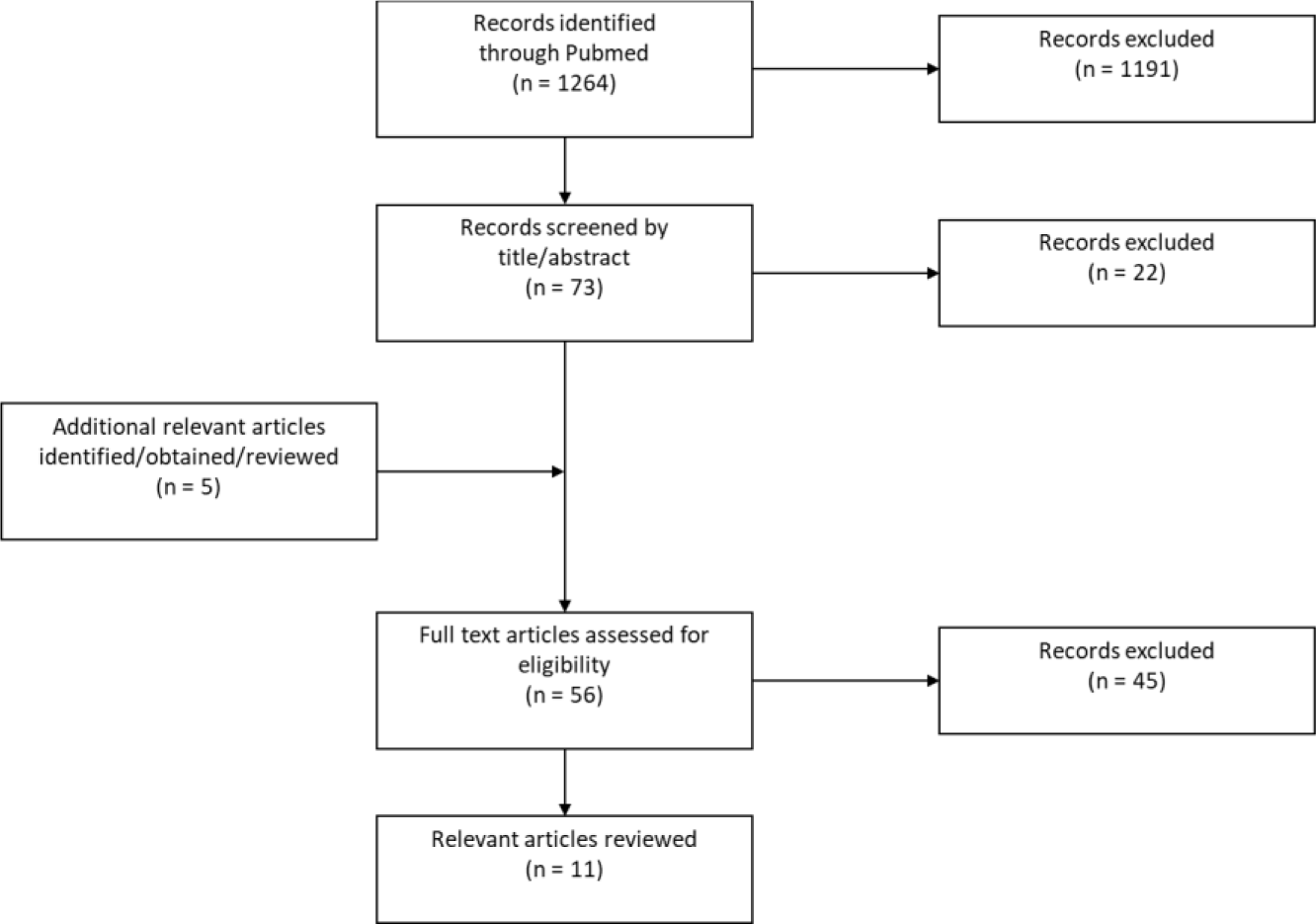
Flow diagram illustrating the literature search process and article selection.

**Table 2.** Inclusion criteria.

|  |  |
| --- | --- |
| - Published in English peer reviewed journals. | - Neuropsychological tests are standardized and have normative data. |
| - ADHD is diagnosed by widely used clinical or research semi-structured or structured interview. | - Results provide diagnostic accuracy statistics, at minimum sensitivity and specificity. |
| - Group study investigation behaviour rating scales for diagnosis, screening, or identification of ADHD. | - Participants are adults, 18 years old or older. |

## Results

### Behavioural rating scales for the screening of ADHD

Two studies focused on distinguishing adults with ADHD from those without the condition, while another two studies aimed to differentiate adults with ADHD from individuals diagnosed with other psychiatric disorders.

The first study used the short version of the WURS (WURS-25) and the ASRS. They found that the WURS-25 had a high level of sensitivity 90% and specificity 88% and the ASRS scale demonstrated a high level of sensitivity of 80% and specificity of 88% in distinguishing between adults who were seeking assessment for ADHD and healthy controls. However there were no mention of PPV nor NPV (Brevik et al., 2020). The second study compared the full version of the WURS with the WURS-25 and found that both versions had high sensitivity and specificity values when distinguishing between control and ADHD groups. Specifically, the full WURS had a sensitivity of 95%, a specificity of 93%, a PPV of 94%, and a NPV of 84%, while the WURS-25 had a sensitivity of 91%, a specificity of 94%, a PPV of 88%, and an NPV of 91%. When differentiating between ADHD and other psychiatric disorders, the full WURS had very high specificity of 94% and NPV of 91% and high sensitivity of 84% and PPV of 88%. In contrast, the WURS-25 demonstrated high specificity of 85% and NPV of 79% and moderate-to-high sensitivity of 62% and PPV of 73% (Gift et al., 2021). Then, Mowlem and colleagues (2019) developed the Mind Excessively Wandering Scale (MEWS) to assess the presence of mind wandering in individuals ADHD. The MEWS defines mind wandering as *“periods in time in which attention switches from a current task to unrelated thoughts and feelings*” (Smallwood & Schooler, 2015). The MEWS is a self-report measure consisting of 12 or 15 items, which are rated on a 4-point Likert-type scale ranging from 0 (indicating “not at all or rarely”) to 3 (indicating “nearly all of the time or constantly”). In their study, the MEWS was found to have a high level of sensitivity of 90% and specificity of 90% in the diagnosis of ADHD using sample of patients with ADHD and healthy controls. The authors reported no PPV nor NPV (Mowlem et al., 2019). Finally, a study was focused on the main scale of the CAARS and the t-scores, which are standardised scores that compares the individual score with the reference group. They found that utilizing a t-score threshold of > 65 for the identification of ADHD was associated with a moderate-to-high sensitivity of 64%, a high specificity of 86%, a moderate PPV of 51%, and a moderate-to-high NPV of 71%. In contrast, employing a t-score threshold of > 70 yielded a low sensitivity of 14%, a very-high specificity of 92%, a moderate PPV of 47%, and a moderate-to-high NPV of 68% (Harrison et al., 2019).

The aforementioned studies presented varying outcomes in comparison to prior research conducted beyond the scope of the reviewed timeframe. For instance, Brevik et al., (2020) found higher specificity (88%) for the ASRS than previously reported by other studies (ranging from 27% to 68%) (Dunlop et al., 2018; Pettersson et al., 2018; Söderström et al., 2014; van de Glind et al., 2013). Similarly, Gift et al., (2021) found a higher specificity for the WURS (93-94%) compared to prior findings (ranging from 57% to 70%) (Luty et al., 2009; McCann et al., 2000). Finally, although the specificity of the CAARS generally remained high across studies, the sensitivity found by Harrison et al. (2019) (64%) was lower than previously reported (97%) when the diagnosis is based on self and observer reports (Luty et al., 2009). These discrepancies between the studies may then be attributed to factors such as differences in sample size, addition of informant’s feedback, the proportion of ADHD subtypes, or whether there was a clinical group. The latter is of particular issue because clinical settings typically involve patients with other psychiatric conditions, such as depression and anxiety, that are known to share comorbidities with ADHD (Anker et al., 2018; Kessler et al., 2006). As a result, there is a higher risk of these patients being erroneously diagnosed.

### Practical recommendations

Our findings yielded the following results, when differentiating between healthy controls and patients with ADHD, the WURS exhibited very-high sensitivity along with the WURS-25 and the MEWS, whereas ASRS had high sensitivity, and the CAARS had moderate sensitivity. Moreover, the WURS and the MEWS showed very-high specificity, while the WURS-25, ASRS and CAARS demonstrated high specificity. The WURS also displayed very-high PPV with a high NPV, whereas the CAARS exhibited moderate PPV with a moderate-to-high. Similarly, when distinguishing between clinical patients and patients with ADHD, the WURS demonstrated very-high sensitivity, and the WURS-25 displayed moderate-to-high sensitivity. Both the WURS and WURS-25 exhibited high specificity. The WURS had a high PPV and a very-high NPV, while the WURS-25 displayed moderate-to-high PPV and high NPV.

Overall, research to date suggest that the CAARS, the MEWS and the WURS are the measures with the best statistical properties for the assessment of adult ADHD (see appendix 2).

### Behavioural Rating scales for the detection of feign ADHD symptoms

A total of seven studies explored indices designed to assess the validity of ADHD symptoms and the potential for feigning results.

To begin, two studies were focused on the validity indexes of the CAARS, which assess for the detection of feigning and non-credible symptom presentation. Becke et al. (2021) created the ADHD credibility index (ACI), a 12-items scale which was then compared with the CII. For the detection of feigning, the ACI had a very high specificity of 98%, a low-to-moderate sensitivity of 30%, and PPV and NPV ranging from 69% to 95% and 58% to 92%, respectively, varying between moderate and very high. Meanwhile, the CII had a very high specificity of 95%, a moderate sensitivity of 46%, and PPV and NPV ranging from 51% to 90% and 63% to 94%, respectively, also varying between moderate and very high (Becke et al., 2021). The authors also created the CII-ACI-Compound Index, a combination of items from both validity indexes. The CII-ACI-Compound Index had a high specificity of 87% to 92%, a moderate sensitivity of 41% to 50%, and PPV ranging from 15% to 87% and NPV ranging from 60% to 97%, varying between low-to-high and moderate-to-high and very high (Becke et al., 2022). Instead of utilising embedded validity indices to detect feigning of ADHD symptoms, Courrégé and colleagues (2019) developed a self-administered measure known as the ADHD Symptom Infrequency Scale (ASIS). Comprising 52 true/false items, the ASIS is subdivided into two scales: the ADHD scale, which includes 19 items designed to align with DSM-5 diagnostic criteria, and the Infrequency scale, which includes 33 items intended to be endorsed more frequently by individuals simulating ADHD than those with a genuine diagnosis. Using a sample composed of patients with diagnosed ADHD and instructed simulators, the infrequency scale of the ASIS demonstrated a high sensitivity of 79% to 86%, specificity of 89%, PPV of 71% to 79%, and a very high NPV of 92% to 93% in the detection of feigning (Courrégé et al., 2019). Another study, which included a clinical control group, reported that the infrequency scale of the ASIS had a very high specificity of 90%, moderate-to-high sensitivity of 71%, moderate-to-high PPV of 65%, and a very high NPV of 93% for detecting feigning (Skeel et al., 2022). Another recent stand-alone measure, the Multidimensional ADHD Rating Scale (MARS), was developed by Potts and colleagues (2021). The MARS includes three categories of items: 18 symptom items, 22 impairment items, and 4 symptom-validity items. Additionally, three “catch” items are incorporated into the assessment to measure effort and attention during the administration. The items are rated on a scale ranging from 0 to 8, where the symptom scale ranges from 0 (“never”) to 8 (“very often”), and the invalid scale ranges from 0 (“not at all”) to 8 (“severe”). For the detection of ADHD, the MARS yielded a high sensitivity of 86% to 92%, a moderate-to-high specificity of 58% to 67%, and PPV ranging from 60% to 65% and high NPV of 86% to 92%. For the detection of feigning, the infrequency scale of the MARS showed a very high specificity of 92%, a moderate-to-high sensitivity of 65%, very high PPV of 92%, and moderate-to-high NPV of 65% (Potts et al., 2021). In another study using a similar sample, the infrequency scale of the MARS and found a specificity of 88%, a moderate-to-high sensitivity of 62%, and moderate-to-high PPV of 63-88% and NPV of 69-87% for the detection of feigning (Potts et al., 2022). Finally, a recent study by Harrison et al. (2022) examined the validity of two new indicators proposed by Aita et al. (2018) for identifying feigned ADHD using the Personality Assessment Inventory (PAI). The PAI is a self-report personality measure consisting of 344 items divided into four scales in which respondents are required to rate each item on a four-point Likert scale ranging from 1 (indicating “false”) to 4 (indicating “very true”) (Morey, 1991). The study utilized a sample of patients with genuine ADHD and healthy controls who were asked to either respond truthfully or feign ADHD. The results revealed that the Item-FAA (Feigned Adult ADHD index using PAI items) had a high specificity of 78%, a moderate sensitivity of 45%, a low PPV of 19%, and a very high NPV of 92%, while the Scale-FAA (Feigned Adult ADHD index using PAI Scales) had a high specificity of 79%, a low-to-moderate sensitivity of 36%, a low PPV of 17%, and a very high NPV of 91% for the detection of feigning (Harrison et al., 2022).

It is noteworthy that the CAARS stands out among behavioural rating scales in that it possesses validity indicators capable of detecting feigned symptoms. In addition to its initial infrequency index, the CII (J. A. Suhr et al., 2011) and the EI (Harrison & Armstrong, 2016), a new index, the ACI, was recently developed (Becke et al., 2021). Although the ACI had very-high sensitivity, its specificity was found to be low-to-moderate. To improve its overall classification accuracy, the authors of the study developed the CII-ACI-Compound, which showed a very high sensitivity but only a moderate specificity (Becke et al., 2022). Although these indexes show promise, further research is necessary to improve their accuracy. Additionally, they are based on different theoretical foundations and may identify various subgroups of examinees as non-credible (Becke et al., 2021). Conversely, as opposed to measures embedded within existing rating scales, some authors have created stand-alone measures that assess ADHD directly while accounting for the existence of feigned symptoms. In spite of the fact that the MARS has shown promise in previous studies (Potts et al., 2021, 2022), its efficacy has not yet been tested on clinical groups, which may have led to inflated results, as explained earlier. On the contrary, the ASIS has exhibited good psychometric properties in its initial validation study (Courrégé et al., 2019) and these findings have already been replicated with a sample of patients reporting symptoms of depression and anxiety (Skeel et al., 2022).

### Practical recommendations

In recent years, there has been a growing emphasis on developing assessment tools aimed at detecting feigned symptoms. This focus arises from the recognition of the inherent subjectivity and lack of specificity in the symptoms evaluated by behavioural rating scales. Current research indicates that stand-alone measures like the ASIS and the MARS exhibit greater accuracy compared to embedded measures. Although these stand-alone measures hold considerable promise, it’s crucial to acknowledge that they are still in the process of refinement and development. Given this evolving landscape, we suggest utilizing the CAARS in conjunction with either the CII or the EI. This recommendation is made with the understanding that further research is necessary to optimize these emerging indices and augment their efficacy.

## Discussion

### Recommendations for research settings

The examination of the existing literature on the use of behavioural rating scales for adult ADHD assessment revealed that they come in various forms and can evaluate different ranges of symptoms (see Figure 2). For example, the WURS primarily assesses childhood symptoms, whereas the CAARS assesses current symptoms and can be used to monitor symptom changes over time. Therefore, we emphasize that the selection of a behavioural rating scale should be based on the primary variable of interest or research objectives. Nonetheless, for a measure that is suitable across a wide range of contexts, we endorse the findings of previous reviews (Marshall et al., 2021; Taylor et al., 2011) and strongly recommend the utilisation of the CAARS. It should be completed collaboratively by both the patient and a pertinent informant, along with either the CII or the EI.

**Figure 2.**
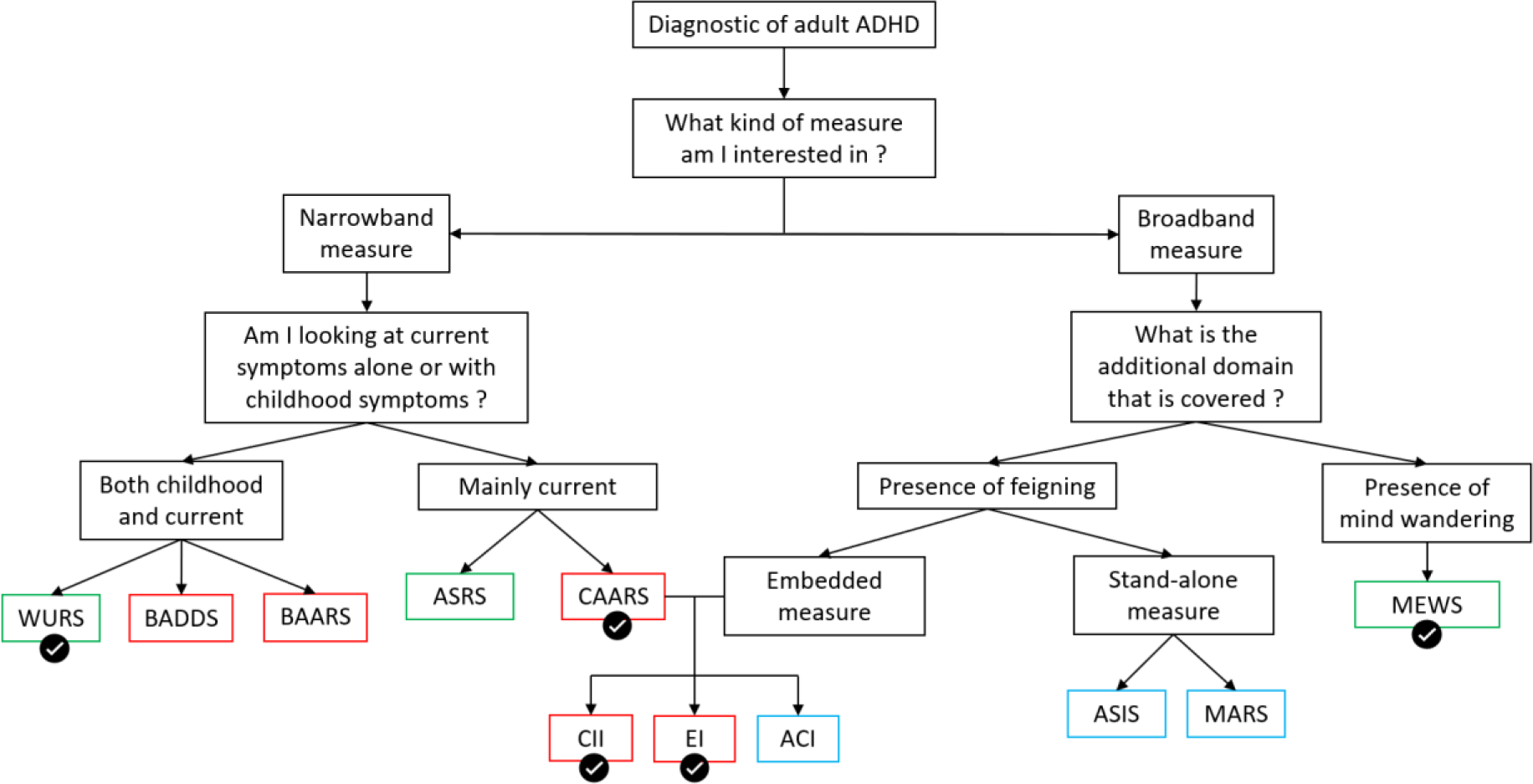
Tree-graph representing behavioural rating scales used in the assessment of adult ADHD. *Note.* The measures are highlighted in green if they are publicly available, in red if they are commercially available and in blue if they are still under development. Note that the CII and EI are in red because they rely on the utilisation of the CAARS. The validation mark emphasizes the rating scales that we recommend based on their accuracy. Narrowband measures refer to scales that focus solely on evaluating specific symptoms associated with ADHD. Broadband measures are assessing a wider range of behaviours. WURS = Wender Utah Rating Scale. ASRS = ADHD self-report scale. CAARS = Conner’s Adult ADHD Rating Scales. CII = CAARS Infrequency Index. ACI = ADHD Credibility Index. ASIS = ADHD Symptom Infrequency Scale. BAARS-IV = Barkley Adult ADHD Rating Scale-Fourth Edition. EI = Exaggeration Index. MEWS = Mind Excessively Wandering Scale. MARS = Multidimensional ADHD Rating Scale.

We propose this scale for the following compelling reasons. Firstly, the literature has identified the CAARS as one of the most accurate measures available. Secondly, the CAARS is a comprehensive tool that assesses a broad range of ADHD symptoms beyond those outlined in the DSM-5 as opposed to the WURS or the ASRS. Thirdly, the CAARS is strengthened as a standalone instrument by its inclusion of validity indicators, which not only enhance its overall utility but also assist in identifying cases where symptoms may have been inaccurately presented. Finally, as previously mentioned, the CAARS is particularly advantageous for studies assessing the efficacy of new ADHD treatments due to its ability to evaluate the current presence and severity of ADHD symptoms. Unlike the WURS, which focuses on historical symptoms, the CAARS can monitor changes in symptoms over time, allowing for a comprehensive assessment of treatment effectiveness. Although the ASIS and MARS are more effective at detecting invalid symptom presentation as well as feigning, they are still underdevelopment and requires independent validation of their effectiveness by other researchers. Accordingly, the validity indicators of the CAARS remain the most optimal choice thus far.

### Strength and limitations

When interpreting the findings of this review, it is important to consider both its strengths and limitations. One potential limitation is that only the Medline database was searched, and the search only retrieved electronic articles. As a result, there’s a possibility of publication bias due to the lack of other sources of information. Another constraint of this paper lies in the instability observed in the base rates of PPV and NPV among the studies reviewed. The variations in these base rates made it challenging to establish a meaningful direct comparison between them and might have led to overestimation of their accuracy. While this paper aimed to update and expand the results of Marshall et al. (2021), we decided not to control for the presence of a clinical group. Therefore, some of the experiments reviewed may not accurately reflect the use of rating scales in a clinical environment, where patients may present other clinical diagnoses that are comorbid with ADHD. Moreover, our decision to apply inclusion criteria that were more flexible than those of Marshall et al. (2021) is a distinct strength of our approach. This choice not only enabled a comprehensive review of a greater number of papers, but also aligned with the research-oriented nature of our focus. Certainly, we recognize that researchers frequently give precedence to evaluating symptoms rather than exclusively concentrating on achieving a diagnosis, as typically seen in clinical contexts. Another strength is our focus on scales detecting feigned or invalid symptom presentation which were often overlooked in prior reviews.

### Conclusion

The objective of this paper was to provide updated insights for researchers by examining recent developments in the use of behavioural rating scales to assess adult ADHD for research purposes. The growing body of research on these scales has raised concerns about their susceptibility to feigning, highlighting the need to consider the objectives of the ADHD assessment when choosing rating scales. While standalone measures may be more appropriate for research settings, combining different measures can enhance diagnostic accuracy in clinical settings. To enhance the practical applicability of feigning measures, further research should involve clinical groups to validate their effectiveness in settings that closely reflects the clinical reality. Future studies should also systematically report sensitivity and specificity along with PPV and NPV to make to facilitate their comparison with existing measures.

### Funding details

The work was supported by the MRC Impact Acceleration Grant to Roi Cohen Kadosh.

### Disclosure statement

R. Mukherjee has delivered presentations for several pharmaceutical companies specializing in ADHD, whereby the funds generated were specifically allocated to the neurodevelopmental teams, with no direct financial compensation received by the author. The remaining authors declare no conflicts of interest.

## Data Availability

All data produced in the present work are contained in the manuscript

## Note

The MARS and the ASIS are still underdevelopment therefore not yet publicly nor commercially available. The WURS and ASRS are publicly available, the MEWS is available without charge by contacting. The ASRS, the CAARS and the WURS are commercially available.

## Appendices

**Appendix 1.**
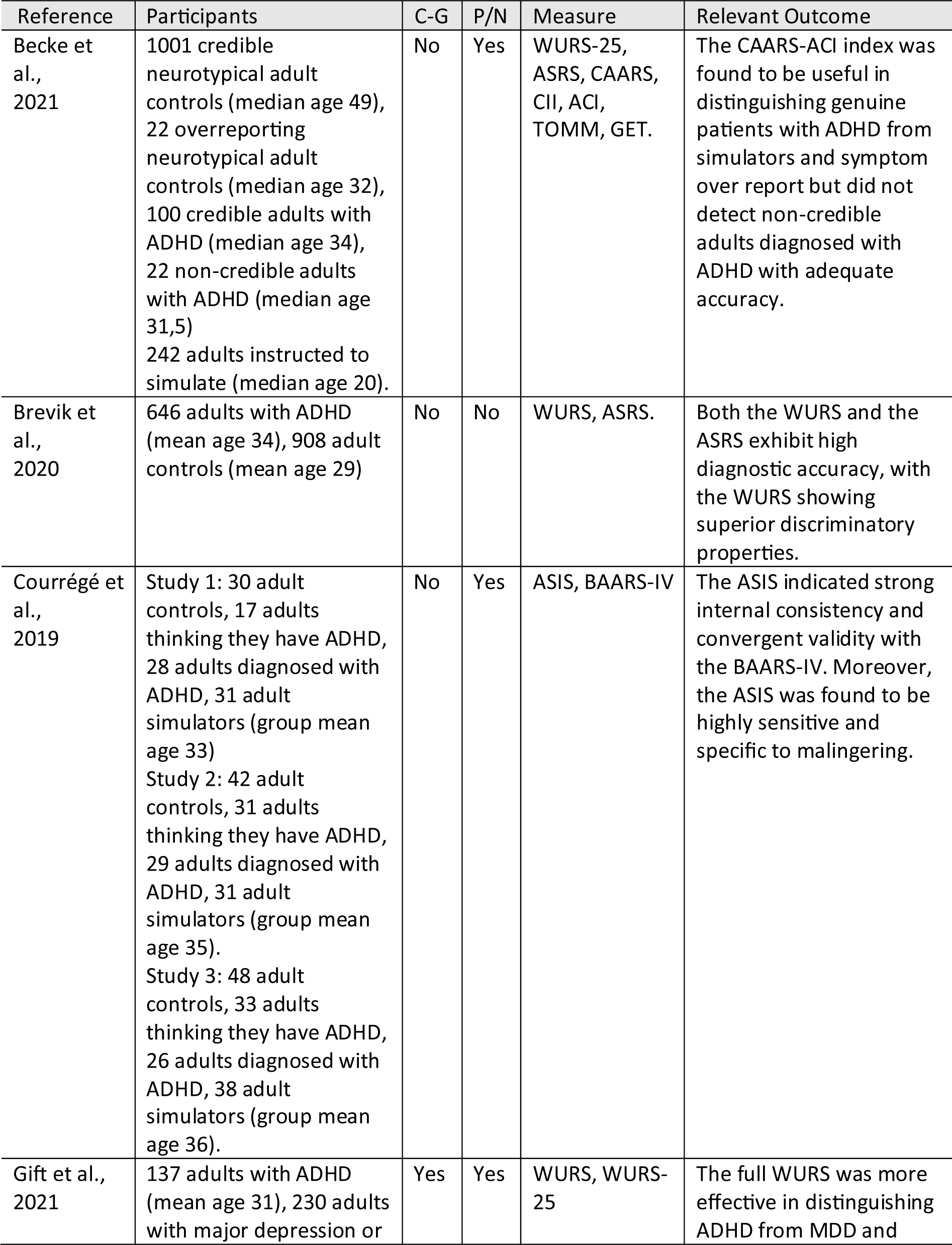
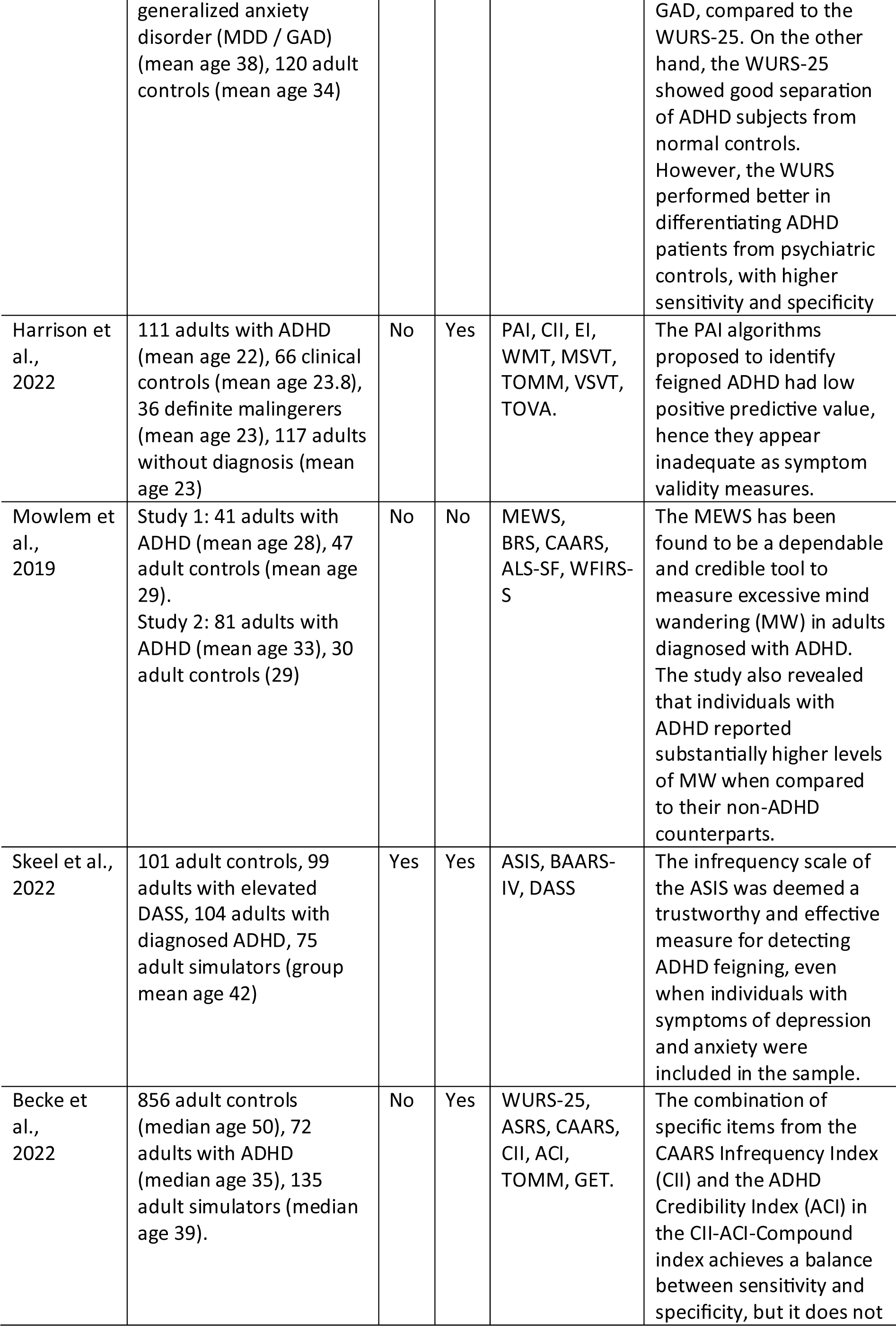
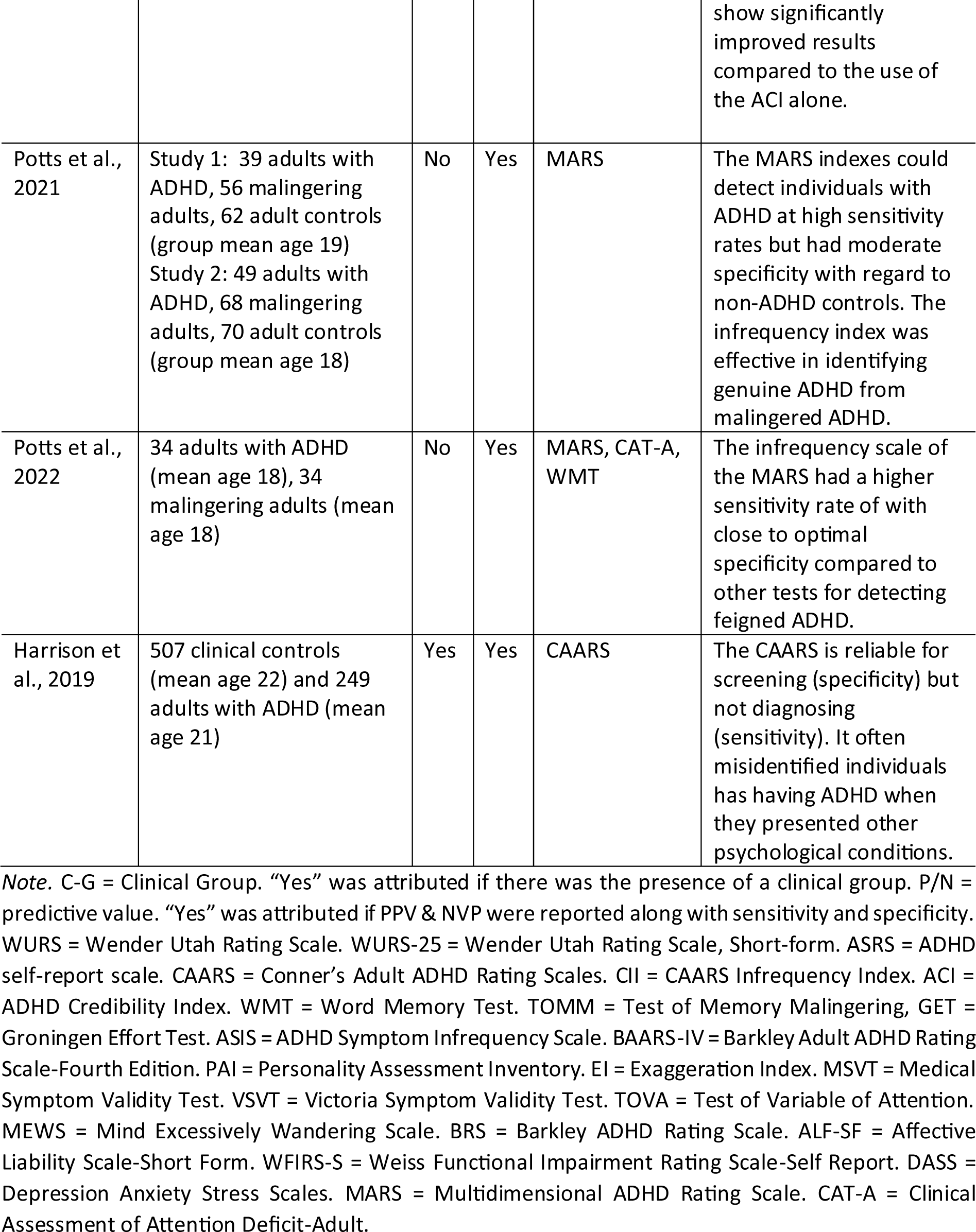
Articles providing diagnostic accuracy for behaviour rating scale used in the assessment of adult ADHD.

**Appendix 2.** Psychometric statistics retrieved from the studies reviewed, arranged by measurement, and shorted by scale.

| Measurement of ADHD |  |  |  |  |  |  |
| --- | --- | --- | --- | --- | --- | --- |
| Scale | Sensitivity % | Specificity % | PPV |  | NPV |  |
|  |  |  | Value % | BR % | Value % | BR % |
| WURS-61 | 84-95 | 93-94 | 88-94 | . | 84-91 | . |
| WURS-25 | 62-90 | 85-94 | 73-88 | . | 79-91 | . |
| ASRS | 80 | 88 | . | . | . | . |
| CAARS | 64 | 86 | 51 | . | 71 | . |
| MARS | 86-92 | 58-67 | 60-65 | 38-40 | 86-92 | 38-40 |
| MEWS | 90 | 90 | . | . | . | . |
| Measurement of Feigning |  |  |  |  |  |  |
| Scale | Sensitivity % | Specificity % | PPV |  | NPV |  |
|  |  |  | Value % | BR % | Value % | BR % |
| ACI | 30 | 98 | 69-95 | 10-50 | 58-92 | 10-50 |
| CII | 46 | 95 | 51-90 | 10-50 | 63-94 | 10-50 |
| ACI-CII | 41 | 87 | 15-87 | 5-50 | 60-67 | 5-50 |
| ASIS | 71-79 | 89-90 | 65-71 | 29 | 92-93 | 29 |
| MARS | 62-65 | 88-92 | 63-92 | 25-50 | 69-87 | 25-50 |
| PAI Item-FAA | 45 | 78 | 19 | 19 | 92 | 19 |
| PAI Scale-FAA | 36 | 79 | 17 | 19 | 91 | 19 |

*Note.* PPV = Positive predictive value. NPV = Negative predictive value. BR = base rate. WURS-61 = Wender Utah Rating Scale, Long form. WURS-25 = Wender Utah Rating Scale, Short-form. ASRS = ADHD self-report scale. CAARS = Conner's Adult ADHD Rating Scales. MARS = Multidimensional ADHD Rating Scale. MEWS = Mind Excessively Wandering Scale. ACI = ADHD Credibility Index. CII = CAARS Infrequency Index. ACI-CII = ACI-CII Compound index. ASIS = ADHD Symptom Infrequency Scale. PAI: Personality Assessment Inventory; Item-FAA: Feigned Adult ADHD index using PAI items; Scale-FAA: Feigned Adult ADHD index using PAI Scales.

## Notes

### Summary of Updates

We have collectively reviewed and incorporated key suggestions, primarily focusing on adjusting the structure to align more closely with a review paper rather than a study paper. Additionally, we have heightened the emphasis on the review's overarching aim.

